# Cumulative incidence of SARS-CoV-2 infection and associated risk factors among frontline health care workers in Paris, France: the SEROCOV prospective cohort study

**DOI:** 10.1101/2021.03.09.21253200

**Authors:** Pierre Hausfater, David Boutolleau, Karine Lacombe, Alexandra Beurton, Margaux Dumont, Jean-Michel Constantin, Jade Ghosn, Alain Combes, Nicolas Cury, Romain Guedj, Michel Djibré, Rudy Bompard, Sandie Mazerand, Valérie Pourcher, Linda Gimeno, Clemence Marois, Elisa Teyssou, Anne-Geneviève Marcelin, David Hajage, Florence Tubach

**Affiliations:** Emergency Department, hôpital Pitié-Salpêtrière, APHP. Sorbonne Université, Paris, France; Sorbonne Université, GRC-14 BIOSFAST, UMR INSERM 1166, IHU ICAN, Sorbonne Université, Paris France; Sorbonne Université, INSERM, Institut Pierre Louis d’Epidémiologie et de Santé Publique (iPLESP), GH AP-HP. Sorbonne Université, Hôpital Pitié-Salpêtrière, Service de Virologie, Paris, France; Infectious Disease Department, Sorbonne Université Hôpital Saint-Antoine, Paris, France; Service de pneumologie - médecine intensive réanimation, hôpital Pitié-Salpêtrière, APHP.Sorbonne Université, Sorbonne Université Inserm UMRS Neurophysiologie respiratoire expérimentale et clinique, Paris, France; Sorbonne University, GRC 29, AP-HP, DMU DREAM, Department of Anaesthesiology and critical care, hôpital Pitié-Salpêtrière, GH APHP.Sorbonne Université, Paris, France; AP-HP, Nord, Service des Maladies Infectieuses et Tropicales, Hôpital Bichat - Claude Bernard, and Université de Paris, INSERM, UMR 1137 IAME, Paris, France; Sorbonne Université, INSERM, UMRS_1166-ICAN, Institute of Cardiometabolism and Nutrition, and Service de médecine intensive-réanimation, Institut de Cardiologie, GH APHP Sorbonne Université Hôpital Pitié–Salpêtrière, Paris, France; Emergency Department, APHP.Sorbonne Université Hôpital Saint-Antoine, Paris, France; Pediatric Emergency Deparment, APHP Hôpital Armand Trousseau– Sorbonne Université, faculté de médecine, Paris, France and Université de Paris, Centre of Research in Epidemiology and Statistics - CRESS, INSERM, F-75004 Paris, France; Service de Médecine intensive Réanimation, APHP.Sorbonne Université Hôpital Tenon, France; Emergency Department, hôpital Tenon, APHP.Sorbonne Université, Paris, France; Service de médecine intensive-réanimation, APHP.Sorbonne Université Hôpital Saint - Antoine, Paris, France; Sorbonne Université, AP-HP, Hôpitaux Universitaires Pitié-Salpêtrière Charles Foix, Service de Maladies infectieuses et Tropicales, INSERM 1136, Institut Pierre Louis d’Epidémiologie et de Santé Publique,75013, Paris, France; APHP.Sorbonne Université, Unité de Recherche Clinique Pitié Salpêtrière Charles Foix, F75013, Paris, France; Unité de Médecine Intensive Réanimation Neurologique, Département de Neurologie, DMU Neurosciences, APHP.Sorbonne Université Hôpital Pitié-Salpêtrière, Paris, France; Sorbonne Université, INSERM, Institut Pierre Louis d’Epidémiologie et de Santé Publique, AP-HP, Sorbonne Université, Hôpital Pitié Salpêtrière, Département de Santé Publique, CIC-1422, F75013, Paris, France

**Keywords:** SARS-CoV-2, serology, seroprevalence, seroconversion, health care worker, smoker

## Abstract

**Background:** With the COVID-19 pandemic, documenting whether health care workers (HCWs) are at increased risk of SARS-CoV-2 contamination and identifying risk factors is of major concern.

**Methods:** In this multicenter prospective cohort study, HCWs from frontline departments were included in March and April 2020 and followed for 3 months. SARS-CoV-2 serology was performed at month 0 (M0), M1, and M3 and RT-PCR in case of symptoms. The primary outcome was laboratory-confirmed SARS-CoV-2 infection at M3. Risk factors of laboratory-confirmed SARS-CoV-2 infection at M3 were identified by multivariate logistic regression.

**Results:** Among 1,062 HCWs (median [interquartile range] age, 33 [28-42] years; 758 [71.4%] women; 321 [30.2%] physicians), the cumulative incidence of SARS-CoV-2 infection at M3 was 14.6% (95% confidence interval [CI] [12.5; 16.9]). Risk factors were the working department specialty, with increased risk for intensive care units (odds ratio 1.80, 95%CI [0.38; 8.58]), emergency departments (3.91 [0.83; 18.43]) and infectious diseases departments (4.22 [0.92; 18.28]); active smoking was associated with reduced risk (0.36 [0.21; 0.63]). Age, sex, professional category, number of years of experience in the job or department, and public transportation use were not significantly associated with laboratory-confirmed SARS-CoV-2 infection at M3.

**Conclusion:** The rate of SARS-CoV-2 infection in frontline HCWs was 14.6% at the end of the first COVID-19 wave in Paris and occurred mainly early. The study argues for an origin of professional in addition to private life contamination and therefore including HCWs in the first-line vaccination target population. It also highlights that smokers were at lower risk.

**Key messages:**

- During the first epidemic wave, 14.6% of 1,062 first-line Health Care Workers had a positive serology and/or RT-PCR test for SARS-CoV-2.
- Most infections occurred early
- Risk was increased by working in infectious diseases (OR 4.22, 95% confidence interval [0.92; 18.28]), emergency (3.91 [0.83; 18.43]) and intensive care units (1.80, [0.38; 8.58])
- Being an active smoker was protective (0.36 [0.21; 0.3]).

## Introduction

The dynamics of the COVID-19 pandemic has deeply affected health services in organizations and potentially exposed healthcare workers (HCWs) to increased risk of SARS-CoV-2 infection. Since the emergence of this new human coronavirus in December 2019 ^1^, knowledge of the modes of transmission has improved weekly, leading to the adaptation of HCWs’ personal protective equipment (PPE). Hence, the risk of SARS-CoV-2 infection among HCWs has been of major concern for frontline institutions, to prevent COVID-19– associated work disruption (at a time when care resources were critical) and HCW-to-patient contamination. In France, SARS-CoV-2–positive RT-PCR tests were first reported in imported cases on January 24, 2020; the generalized lockdown began on March 17 and emergency room visits for possible COVID-19 peaked on week 13, decreasing thereafter, which led the French government to ease lockdown restrictions on May 11.

Although some cross-sectional studies have reported SARS-CoV-2 seroprevalence among HCWs being close to that of the general population, ^2–7^ little is known about the dynamics of SARS-CoV-2 contamination related to the epidemic waves and the risk factors in this population.

In the ascendant phase of the first COVID-19 wave in France, we initiated a multicenter prospective cohort study to estimate the cumulative incidence of laboratory-confirmed SARS-CoV-2 infection over the early phase of this outbreak among frontline public hospital HCWs in Paris and identify the risk factors (SEROCOV, ClinicalTrials.gov NCT04304690).

## Patients and methods

### Setting, design and participants

The SEROCOV multicenter prospective cohort study was conducted in 4 adult hospitals (Bichat, Pitié-Salpêtrière, Saint-Antoine, Tenon) and 1 pediatric hospital (Trousseau) of the Assistance Publique-Hôpitaux de Paris (AP-HP) network. Participants were included from March 16 to April 24 2020, with a 3-month follow-up. Bichat and Pitié-Salpêtrière hospitals are dedicated referent hospitals for emerging biological risk at AP-HP and primarily received all COVID-19 confirmed cases, from the epidemic onset in Paris until mid-March 2020, when all AP-HP hospitals were positioned to take care of COVID-19 patients.

The SEROCOV study was approved by the ethics committee (CPP Sud-Ouest et Outre-Mer I, approval no. 2-20-023 id7257) and all participants signed informed consent before inclusion.

All medical or paramedical staff who worked in COVID-19 frontline departments (emergency, infectious diseases, intensive care units and virology laboratory) of the selected hospitals were informed about the study and asked to participate. Staff who were not active during the inclusion period were not eligible. After signing informed consent, each participant underwent venous blood sampling for SARS-CoV-2 serology and was asked to complete a self-reported questionnaire on baseline characteristics and pre-inclusion symptoms suggestive of COVID-19. Thereafter and up to 3 months after inclusion, the HCW participants received a weekly reminder to repeat the survey on symptoms and, in case of symptoms, asked for a nasopharyngeal swab for SARS-CoV-2 RT-PCR. At month 1 (M1) and M3 after inclusion, venous blood sampling was again systematically proposed for serology testing.

### Anti–SARS-CoV-2 IgG assay for serology testing

Blood samples were sent immediately to the virology department of each participating hospital and centrifuged, and EDTA plasma was frozen at −20°C until tested by batch in the Pitié-Salpêtrière virology laboratory. Plasma samples were analyzed on the Abbott Architect platform with the Abbott SARS-CoV-2 IgG assay, targeting the viral nucleoprotein.^8^ Results were interpreted as recommended by the manufacturer: an index value < 0.5 was considered negative, 0.5 to 1.4 weak positive, and ≥1.4 positive. When SARS-CoV-2 infection was suspected in an HCW during the study period, the COVID-19 diagnosis was established by RT-PCR on a nasopharyngeal swab with the Cobas SARS-CoV-2 kit (Roche Diagnostics) or RealStar SARS-CoV-2 RT-PCR kit (Altona), as previously reported. ^9^

### Outcomes

The primary outcome was laboratory-confirmed SARS-CoV-2 infection at M3 (i.e., positive serology results [positive or weak positive result on SARS-CoV-2 IgG assay] and/or positive RT-PCR result on a nasopharyngeal swab). Secondary outcomes were positive serology for SARS-CoV-2 (considering weak positive serology as positive) at M0, M1 and M3.

### Covariates

Baseline characteristics collected were demographics, smoking status (active, former or non-smoker), public transportation use, and job-related data (hospital, type of ward [emergency, infectious diseases, intensive care unit and virology laboratory]), professional category, number of years of experience in the job or department, and night or day shift. Physicians (senior or medical students, including residents), nurses and care assistants were considered high-risk HCWs.

At baseline and up to 3 months after inclusion, participants completed a self-reported questionnaire weekly on symptoms suggestive of COVID-19 and as appropriate, SARS-CoV-2 RT-PCR results, the diagnosis retained, and COVID-related sick days and/or hospitalization.

At M3, adherence to PPE recommendations during the study period was reported on a 5-point Likert scale (from “never” to “systematically”).

### Statistical analysis

The number of eligible HCWs in participating services was estimated at approximately 1,000 at the time this study was designed. This planned sample size allowed for estimating a cumulative incidence of laboratory-confirmed SARS-CoV-2 infection at M3 of 5% (95% confidence interval [CI] [3.7; 6.5]), a cumulative incidence of 10% (8.2; 12), and a cumulative incidence of 20% (17.6; 22.6).

Staff characteristics are reported as percentages for categorical variables and median (interquartile range [IQR]) for continuous variables.

Multiple imputation was used to replace missing serology testing at M0, M1 or M3. Briefly, 10 copies of the dataset were created with the missing values replaced by imputed values based on observed data including observed serology results and characteristics of participants (including age, sex, job-related data, smoking status, public transportation use, adherence to PPE recommendations and occurrence of ageusia and/or anosmia over the study period). Each dataset was then analyzed by using standard statistical methods, and the results from each dataset using the statistical methods described below were pooled into a final result by using Rubin’s rule. ^10^

For binomial proportions, confidence intervals were estimated by the Wilson method for multiple imputation. ^11^

Risk factors of laboratory-confirmed SARS-CoV-2 infection at M3 were assessed in the whole cohort by univariate and multivariate logistic regression. Baseline variables included in the multivariate model were defined a priori (age, sex, job-related data, public transportation use and smoking status), and no variable selection was performed. A second logistic model was developed for the same outcome among high-risk HCWs only (i.e., excluding virology staff and jobs rated as “other”) and accounting for adherence to PPE recommendations. Finally, risk factors of laboratory-confirmed SARS-CoV-2 infection at inclusion (M0) were evaluated by the same approach.

All the analyses were computed at a two-sided α level of 5% with R software, version 4.0.0.

## Results

From March 16 to April 24, 2020, 1,177 HCWs were included; 115 were excluded because they did not work in the participating departments, so 1,062 HCWs remained for analysis. The participation rate was 77% for medical staff and 59.5% for paramedical staff. The main characteristics of the study population are in Table 1. Median [IQR] age was 33 [28-42] years, 758 (71.4%) were women, 741 (69.8%) were paramedical staff (including 405 [38.1%] nurses) and 321 (30.2%) were physicians (including students); 380 (35.8%) were working in an emergency department, 282 (26.6%) in an infectious diseases unit, 355 (33.4%) in an intensive care unit and 45 (4.2%) in a virology laboratory. Among the participants, 250 (26.9%) reported actively smoking, and 387 (36.4%) used public transportation.

**Table 1:** Baseline characteristics of the study population according to SARS-CoV-2 serology at inclusion.

|  | **Negative** | **Positive** | **Total\*** |
| --- | --- | --- | --- |
|  | **SARS-CoV-2** | **SARS-CoV-2** | **(N=1062)** |
|  | **serology** | **serology** |  |
|  | **(N=999)** | **(N=61)** |  |
| Age | 33 [28-42.5] | 31 [26-38] | 33 [28-42] |
| Women | 717 (71.8%) | 40 (65.6%) | 758 (71.4%) |
| Professional category |  |  |  |
| Senior physicians | 208 (20.8%) | 12 (19,7%) | 221 (20.8%) |
| Medical students (including residents) | 89 (8.9%) | 11. (18%) | 100 (9.4%) |
| Care assistants | 197 (19.7%) | 6 (9.8%) | 203 (19.1%) |
| Nurses | 378 (37.8%) | 27 (44.7%) | 405 (38.1%) |
| Others | 127 (12.7%) | 5 (8.2%) | 133 (12.5%) |
| Working department |  |  |  |
| Emergency department | 372 (37.2%) | 7 (11.5%) | 380 (35.8%) |
| Infectious diseases unit | 239 (23.9%) | 43 (70.5%) | 282 (26.6%) |
| Intensive care unit | 343 (34.3%) | 11 (18%) | 355 (33.4%) |
| Virology laboratory | 45 (4.5%) | 0 (0%) | 45 (4.2%) |
| Working in a COVID-free unit | 80 (8%) | 2 (3.3%) | 82 (7.7%) |
| Working in a referent hospital for emerging biological risk | 519 (52%) | 32 (52.5%) | 552 (52%) |
| Night shift | 123 (12.3%) | 5 (8.2%) | 129 (12.2%) |
| Missing data, No | 1 | 0 | 1 |
| Experience in the job (months) | 84 [36-180] | 72 [30-168] | 84 [36-180] |
| Missing data, No | 5 | 0 | 5 |
| Experience in the department (months) | 36 [10-84] | 24 [6-72] | 36 [9-84] |
| Missing data, No | 14 | 0 | 14 |
| Public transportation use | 369 (40.5%) | 18 (40.9%) | 387 (40.5%) |
| Missing data, No | 88 | 17 | 105 |
| Active smoker | 246 (27.8%) | 4 (8.9%) | 250 (26.9%) |
| Missing data, No | 115 | 16 | 132 |
| Symptoms suggestive of COVID-19 before inclusion | 125 (13.3%) | 29 (50%) | 154 (15.4%) |
| Missing data, No | 57 | 3 |  |
| If yes: Fever | 29 (23.2%) | 13 (44.8%) | 42 (27.3%) |
| Anosmia and/or agueusia | 10 (8%) | 14 (48.3%) | 24 (15.6%) |
| Respiratory symptoms | 56 (44.8%) | 13 (44.8%) | 69 (44.8%) |
| Gastrointestinal symptoms | 25 (20%) | 2 (6.9%) | 27 (17.5%) |
| Adherence to PPE recommendations over the study period** |  |  |  |
| Wear a surgical mask | 829 (93%) | 40 (85%) | 870 (93.1%) |
| Wear an FFP2 mask to take nasopharyngeal swabs | 521 (78%) | 28 (82%) | 549 (77.8%) |
| Wear an N95 mask to handle a confirmed COVID-19 case | 578 (74%) | 24 (56%) | 603 (73.2%) |
| If wearing a mask (surgical or FFP2), change every 4 hr | 570 (65%) | 22 (47%) | 592 (63.9%) |
| Hydro-alcoholic friction or hand washing before and after each patient contact | 831 (99%) | 45 (98%) | 877 (98.5%) |

Data are n (%) for qualitative variables and median [interquartile range] for quantitative variables
\* HCWs with positive, negative or missing (n=2) serology at inclusion, thus 2 more individuals than the sum of the 2 other columns.
\*\*Collected at the end of the study on a 5-point Likert scale (0, never, to 5, systematically), reported as the proportion of HCWs rating 4 or 5; PPE: personal protective equipment

Overall, 1,060 (99,8%) participants underwent baseline serology testing at inclusion, 1,004 (94.5%) at M1, and 938 (88.3%) at M3; 903 (85%) had a SARS-CoV-2 serology result available for the 3 samples (Figure 1). At M3, during the whole study period, 93.0% of HCWs reported that they wore a surgical mask, 77.8% wore an N95 mask when performing high-risk tasks (nasopharyngeal swab), and 98.5% washed hands and/or used hydro-alcoholic hand friction before and after each contact with a patient (table 1). The dates of inclusion and follow-up serologies according to the epidemic curve are in Figure 2.

**Figure 1:**
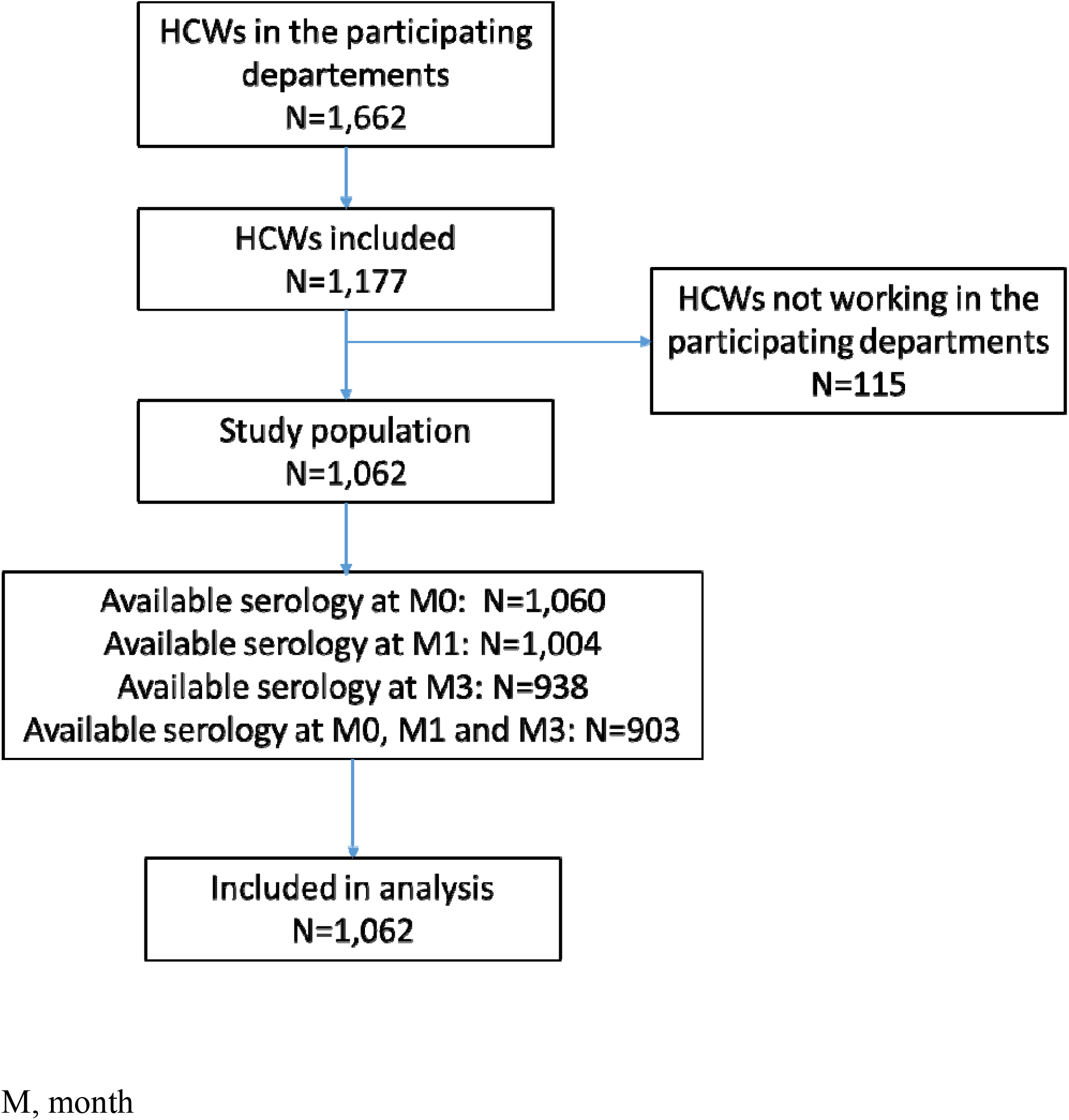
Flow chart.

**Figure 2:**
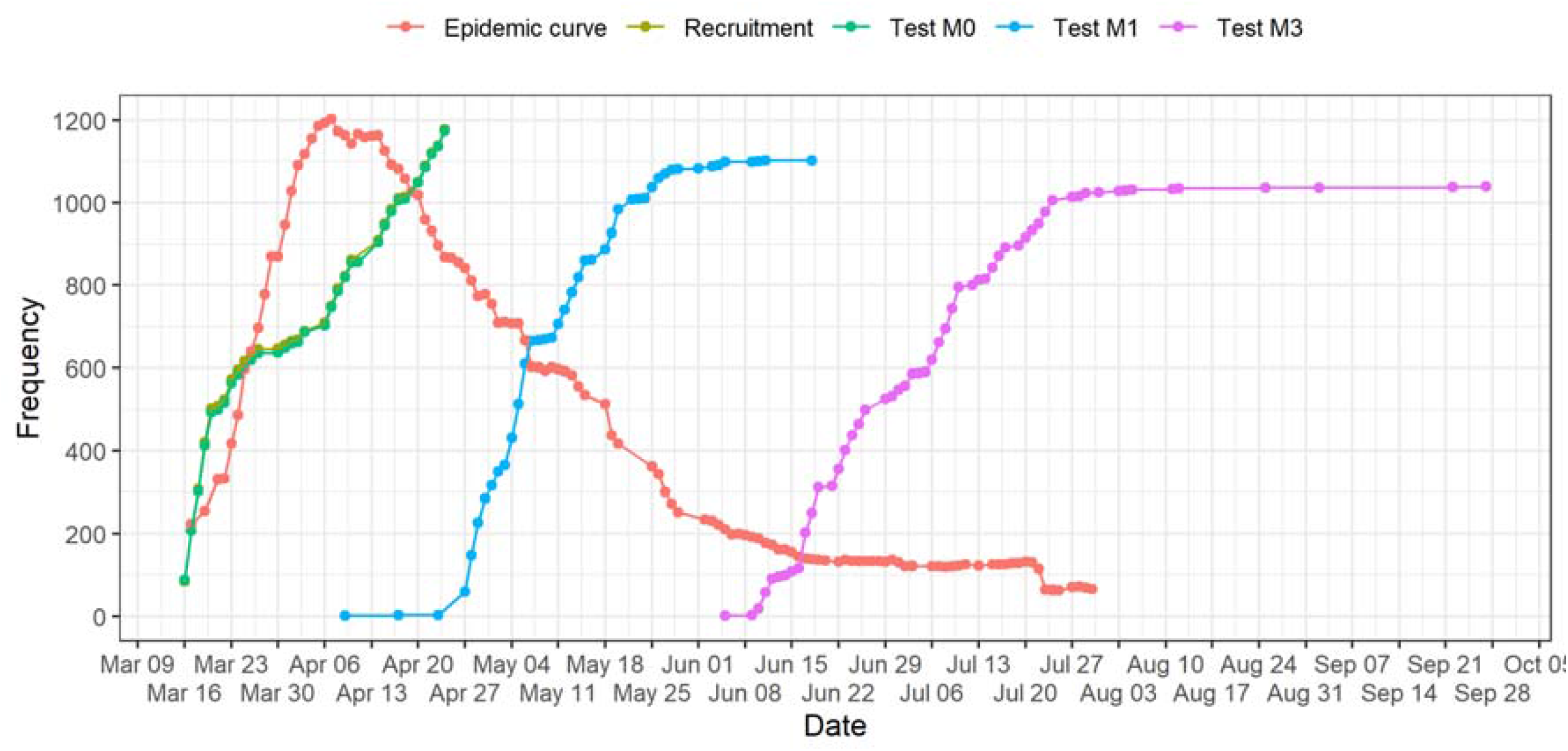
Dates of inclusion (recruitment) and serology tests according to the epidemic curve (inpatients with documented COVID-19 in AP-HP hospital network)

At baseline, 154/1062 (15.4%) HCWs with available data reported having symptoms suggestive of COVID-19 before inclusion, mainly fever (n=42), myalgia (n=34), headache (n=54), ageusia and/or anosmia (n=24), cough (n=55), respiratory signs (n=69), and gastrointestinal signs (n=27) (Table 1).

At inclusion, 61 (5.8%) HCWs had positive SARS-CoV-2 serology results: 23 (7.2%) physicians and 38 (5.1%) paramedical staff. Reports of previous symptoms suggestive of COVID-19 were more frequent for HCWs with positive than negative serology at inclusion (50% vs 13.3%; p<0.0001), particularly symptoms very suggestive of COVID-19 such as ageusia/anosmia (48.3% vs 8%). Conversely, 50% of participants with positive serology at inclusion reported no previous symptoms.

Because of symptoms suggestive of COVID-19, 33 participants submitted an intermediate nasopharyngeal swab for SARS-CoV-2 RT-PCR between inclusion and M3: 14 (42.4%) had a positive result, 2 with negative results at all serology points. Among the 134 participants with seropositive results at inclusion or M1, 11 (8.2%) had negative serology results at M3, including 8 with positive results at inclusion.

### Outcomes

The estimated cumulative incidence of laboratory-confirmed SARS-CoV-2 infection at M3 after imputation of missing data was 14.6% (95%CI [12.5; 16.9]), and the difference across professional categories was not statistically significant, although the incidence was highest for medical students (23.5% [20.8; 26.4]) and higher in nurses than in care assistants (Table 2).

**Table 2:** Laboratory-confirmed infection with SARS-CoV-2 at month 3 (M3) and seroprevalence at M0, M1 and M3 after imputation of missing data, according to professional category

|  | All | Senior physicians | Medical students | Care assistants | Nurses | Others | p value* |
| --- | --- | --- | --- | --- | --- | --- | --- |
| Serology positive |  |  |  |  |  |  |  |
| at M0 | 5.9%<br>[4.7 ; 7.5] | 6.3%<br>[5.0 ; 7.9] | 12.9%<br>[11.0 ; 15.1] | 3.9%<br>[2.9 ; 5.2] | 7.1%<br>[5.7 ; 8.9] | 5.3%<br>[4.1 ; 6.8] | 0.06 |
| at M1 | 12.9%<br>[10.9 ; 15.1] | 11.8%<br>[9.9 ; 13.9] | 21.2%<br>[18.7 ; 23.8] | 11.6%<br>[9.8 ; 13.8] | 14.9%<br>[12.9 ; 17.2] | 10.5%<br>[8.8 ; 12.5] | 0.13 |
| at M3 | 13.0%<br>[11.1 ; 15.2] | 12.1%<br>[10.3 ; 14.3] | 20.4%<br>[18.7 ; 23.2] | 11.6%<br>[9.7%; 13.8] | 14.8%<br>[12.8 ; 17.1] | 10.5%<br>[8.8 ; 12.5%] | 0.21 |
| Laboratory-confirmed SARS-CoV-2 infection at M3 | 14.6%<br>[12.5 ; 16.9] | 14.2%<br>[12.2 ; 16.5] | 23.5%<br>[20.8 ; 26.4] | 12.5%<br>[10.6 ; 14.7] | 16.6%<br>[14.4 ; 18.9] | 11.3%<br>[9.5 ; 13.4] | 0.12 |

Data are % (95% confidence interval).
\*Comparison across professional categories

The estimated seroprevalence after imputation of missing data at M0, M1, and M3 was 5.9% [4.7; 7.5], 12.9% [10.9; 15.1] and 13.0% [11.1; 15.2], with the same trend across professional categories but not statistically significant.

Of the 147 participants with laboratory-confirmed SARS-CoV-2 infection at M3, only 88 (59.9%, 95% CI [51.47; 67.85]) reported symptoms suggestive of COVID-19 before or during the study period. Among the 416 participants with negative serology at M0 and reporting symptoms suggestive of COVID-19 during the study period, in those with and without documented SARS-CoV-2 infection, 58.7% vs 13.3% reported sick leave related to these symptoms (p<0.0001). Among the 147 participants with laboratory-confirmed infection by SARS-CoV-2 at M3, 3 were hospitalized for COVID-19 and none died.

### Risk factors for SARS-CoV-2 positive serology at inclusion

After multiple imputation for missing data, on multivariate analysis (Table 3), risk factors for SARS-CoV-2 positive serology at inclusion (i.e., during the increasing phase of first epidemic wave) were the department where the participant worked, with increased risk for intensive care units (OR 1.29, 95%CI [0.47; 3.51]) and particularly infectious disease units (6.61 [2.64; 16.54]) versus emergency departments (virology laboratory was not included because no virology staff were positive at inclusion). Being an active smoker reduced the risk (0.28 [0.10; 0.82]). Age, sex, using public transportation, professional category, working in a referent hospital for emerging biological risk, number of years of experience in the job or department, and night shift were not significantly associated with baseline positive serology.

**Table 3:** Risk factors of laboratory-confirmed SARS-CoV-2 infection at inclusion (after imputation of missing data; N=1,017*)

|  | Univariate analysis |  |  | Multivariate analysis |  |  |
| --- | --- | --- | --- | --- | --- | --- |
|  | OR | [95% CI] | p value | OR | [95% CI] | p value |
| Age | 0.99 | [0.96; 1.02] | 0.42 | 0.98 | [0.95; 1.01] | 0.26 |
| Women | 0.76 | [0.44; 1.32] | 0.33 | 0.60 | [0.30; 1.23] | 0.16 |
| Working in a referent hospital for emerging biological risk | 1.11 | [0.66; 1.87] | 0.69 | 1.08 | [0.56; 2.09] | 0.83 |
| Working department |  |  | <0.0001 |  |  | <0.0001 |
| Emergency department | 1 |  |  | 1 |  |  |
| Intensive care unit | 1.68 | [0.64; 4.39] |  | 1.29 | [0.47; 3.51] |  |
| Infectious diseases department | 9.46 | [4.18; 21.40] |  | 6.61 | [2.64; 16.54] |  |
| Professional category |  |  | 0.08 |  |  | 0.50 |
| Physicians | 1 |  |  | 1 |  |  |
| Medical students | 2.17 | [0.92; 5.11] |  | 1.62 | [0.42; 6.28] |  |
| Care assistants | 0.51 | [0.19; 1.40] |  | 1.18 | [0.38; 3.63] |  |
| Nurses | 1.20 | [0.59; 2.41] |  | 1.86 | [0.77; 4.50] |  |
| Nurses | 0.89 | [0.30; 2.60] |  | 0.78 | [0.22; 2.69] |  |
| Experience in the department ≥12 months | 0.71 | [0.41; 1.23] | 0.22 | 1.27 | [0.49; 3.25] | 0.62 |
| Experience in the job ≥12 months | 1.20 | [0.47; 3.07] | 0.71 | 0.94 | [0.25; 3.56] | 0.92 |
| Night shift | 0.61 | [0.24; 1.56] | 0.30 | 0.52 | [0.12; 2.31] | 0.39 |
| Public transportation use | 1.11 | [0.62; 2.02] | 0.65 | 0.99 | [0.51; 1.94] | 0.84 |
| Active smoker | 0.25 | [0.088; 0.701] | 0.009 | 0.28 | [0.10; 0.82] | 0.02 |

OR: odds ratio; CI: confidence interval
\*HCWs from virology department were excluded because none had a positive serology result at inclusion

### Risk factors for laboratory-confirmed SARS-CoV-2 infection at M3

After multiple imputation for missing data, on multivariate analysis (Table 4), risk factors for laboratory-confirmed SARS-CoV-2 infection at M3 (i.e., at the end of the first epidemic wave) were the department where the participant worked, with increased risk for intensive care units (OR 1.80, 95%CI [0.38; 8.58]) and particularly emergency departments (3.91 [0.83; 18.43]) and infectious diseases units (4.22 [0.92; 18.28]) versus the virology laboratory. Being an active smoker reduced the risk (0.36 [0.21; 0.3]). Age, sex, use of public transportation, professional category, working in a referent hospital for emerging biological risk, number of years of experience in the job or department, or night shift were not significantly associated with laboratory-confirmed SARS-CoV-2 infection at M3.

**Table 4:** Risk factors of laboratory-confirmed SARS-CoV-2 infection at M3 (after imputation of missing data; N=1,062))

|  | Bivariate analysis |  |  | Multivariable analysis |  |  |
| --- | --- | --- | --- | --- | --- | --- |
|  | OR | [95% CI] | p value | OR | [95% CI] | p value |
| Age | 0.98 | [0.97; 1.00] | 0.06 | 0.99 | [0.97; 1.01] | 0.41 |
| Women | 1.05 | [0.70; 1.56] | 0.79 | 1.04 | [0.65; 1.67] | 0.85 |
| Working in a referent hospital for emerging biological risk | 0.90 | [0.63; 1.28] | 0.57 | 1.16 | [0.76; 1.77] | 0.49 |
| Working department |  |  | 0.0002 |  |  | 0.003 |
| Virology laboratory | 1 |  |  | 1 |  |  |
| Intensive care unit | 2.07 | [0.47; 9.02] |  | 1.80 | [0.38; 8.58] |  |
| Emergency department | 4.10 | [0.97; 17.39] |  | 3.91 | [0.83; 18.43] |  |
| Infectious diseases department | 5.65 | [1.33; 23.09] |  | 4.22 | [0.92; 18.28] |  |
| Professional category |  |  | 0.12 |  |  | 0.38 |
| Senior physicians | 1 |  |  | 1 |  |  |
| Medical students | 1.79 | [0.93; 3.44] |  | 1.69 | [0.68; 4.18] |  |
| Care assistants | 0.85 | [0.47; 1.53] |  | 0.99 | [0.50; 1.96] |  |
| Nurses | 1.24 | [0.77; 2.00] |  | 1.49 | [0.85; 2.63] |  |
| Others | 0.71 | [0.35; 1.42] |  | 0.87 | [0.40; 1.87] |  |
| Experience in the department $\geq 12$ months | 0.78 | [0.53; 1.16] | 0.23 | 1.14 | [0.60; 2.17] | 0.70 |
| Experience in the job $\geq 12$ months | 0.86 | [0.48; 1.55] | 0.63 | 0.97 | [0.42; 2.24] | 0.83 |
| Night shift | 1.03 | [0.60; 1.75] | 0.88 | 1.00 | [0.52; 1.90] | 0.91 |
| Public transportation use | 1.21 | [0.84; 1.75] | 0.32 | 1.17 | [0.79; 1.75] | 0.44 |
| Active smoker | 0.38 | [0.22; 0.66] | 0.0005 | 0.36 | [0.21; 0.63] | 0.0003 |

OR: odds ratio; CI: confidence interval

In the subgroup of high-risk HCWs (i.e., physicians, nurses and care assistants in clinical wards), adjustment on adherence to PPE recommendations did not change these results (Supplementary Table 5).

## Discussion

We conducted this multicenter prospective cohort study among frontline HCWs to investigate SARS-CoV-2 infection in HCWs during the first COVID-19 epidemic wave in Paris, France. We report a 5.8% baseline (early phase) SARS-CoV-2 seroprevalence rate together with a 3-month 14.6% rate of laboratory-confirmed SARS-CoV-2 infection (end of first wave). Most infections occurred very early, that is, diagnosed at M1. Among the identified risk factors was the working department, infectious disease units being at highest risk at the very early phase, then emergency departments during the epidemic peak, with the same level of risk for both at the end of the wave and intensive care units with an intermediate risk, with the virology laboratory staff as reference. Active smokers were at reduced risk. Demographics, other job-related characteristics, adherence to PPE, and use of public transportation were not significantly associated with SARS-CoV-2 infection. To our knowledge, our study is the largest reporting the longitudinal evolution of anti–SARS-CoV-2 antibodies in HCWs over a 3-month period.

The seroprevalence in the general population estimated in the SAPRIS study^12^ in the same geographical area (Greater Paris) in May 2020, corresponding to M1 in the SEROCOV study, was significantly lower than in the SEROCOV first-line HCWs: 6.4% in the 20-to 59-year-old population of the SAPRIS study (using a similar anti-nucleoprotein assay and considering weak positive as positive as in SEROCOV) versus 13.3% in SEROCOV (*p*<0.0001). Previous published HCW surveys reported seroprevalence ranging from 3.4% to 13.7%. However, the heterogeneity in type of HCW tested, type of assay used, period of testing and countries with incomparable SARS-CoV-2 epidemic burden precludes a face-to-face comparison. In a multicenter cross-sectional study in the New York city area, Moscola et al. reported a 13.7% prevalence (95%CI [13.4; 14.0]) of SARS-CoV-2 antibodies in 40,329 HCWs, a rate similar to that among adults randomly tested in New York state (14.0%). ^3^ However, in another cross-sectional study in Roslyn, New York, in one hospital, employees had a significantly lower serology-positive rate than the general population (9.9% vs 16.7%, *P* < 0.001).^4^ Stubblefield et al. reported a 7.6% seroprevalence among 249 frontline HCWs during the first month of epidemic in Nashville, Tennessee, whereas Jespersen et al. reported a 3.4% seroprevalence (95% CI [2.5; 3.8]) among 17,971 HCWs, administrative personnel, pre-hospital services and specialist practitioner clinics in Denmark, with high geographical seroprevalence variation (from 1.2% to 11.9%). ^6,7^ In the United States, in a Multistate Hospital Network, Self et al. reported 6.0% positive serology in front-line HCWs.^5^ Among the 194 participants with positive results, 56 (29%) reported no symptoms in the previous weeks. In our study, this asymptomatic proportion was 50% among participants with positive serology at inclusion. Conversely, we confirm that anosmia and/or ageusia during the epidemic wave was a discriminative clinical sign, although with low sensitivity. This result confirms the Belgian monocentric study findings reporting of anosmia associated with an OR for positive serology of 7.78 ([95%CI [5.22; 11.53]). ^2^

Most HCW contaminations occurred very early during the epidemic wave, almost half documented at baseline (positive serology: 5.9%, 95% CI [4.7; 7.5]) and 12.9% at M1 (corresponds to infections during the ascendant phase of the epidemic wave as shown in Figure 2). The kinetics of these infections probably reflects both infections in the private sphere, following the epidemic curve of the general population, but also the effectiveness of better adherence to protective measures in professional activity related to an enforcement of PPE and standard measures as the epidemic progressed. The family and friends or professional origin of HCW contamination is still debated.^13^ In a single center study in Belgium of 3,056 HCWs (tested with an IgG/IgM rapid lateral flow assay), the OR for positive serology was 3.15 (95% CI [2.33; 4.25]) when reporting household contact with a suspected or confirmed COVID-19 case. ^2^ Conversely, a large study in Denmark of 17,971 hospital staff (based on the adjusted seroprevalence according to living and working places) found risk of SARS-CoV-2 positive serology associated with workplace rather than place of living. ^7^

Recently, a large survey of HCWs in the United Kingdom reported anti-spike or anti-nucleocapsid IgG antibodies associated with substantially reduced risk of SARS-CoV-2 reinfection and highlighted that individual HCWs middle-/long-term protection by post-COVID self-immunization is a major concern.^14^ Our data support the persistence of SARS-CoV-2 antibodies at 3 months: only 11 (8%) participants with positive serology had later negative results. Controversial results were reported by Patel et al. for 249 HCWs with a positive serology rate decreasing from 7.6% at baseline to 3.2% at 60 days, whereas Gudbjartsson et al. showed no decline in anti–SARS-CoV-2 antibodies at 4 months after COVID-19 diagnosis. ^15,16^

Workers in first-line clinical departments (infectious diseases, emergency, intensive care unit) were at higher risk of laboratory-confirmed SARS-CoV-2 infection as compared with virology laboratory staff (Table 3). In a large study in Denmark, Jespersen et al. reported the highest adjusted seroprevalence in emergency departments: 29.7% (95% CI [23.1; 37.6]); departments with no or limited patient contact had the lowest seroprevalence: 1.79% (0.31; 3.90).^7^ From January to mid-March 2020, infectious disease departments and intensive care units directly admitted patients with suspected COVID-19 from home or the general practitioner’s office, bypassing the emergency department, and were therefore initially more exposed to SARS-CoV-2 than emergency department workers. This situation may explain the association of SARS-CoV-2 positive serology with working in intensive care units (OR 1.2 [95%CI (0.47; 3.51)]) and especially infectious diseases units (6.61, [2.64; 16.54]) at inclusion versus emergency departments (Table 3). Virology laboratory staff, although handling numerous SARS-CoV-2–contaminated samples, are probably more concerned and stricter about PPE enforcement to prevent contamination and had no staff with positive serology at inclusion, together with the lowest seroprevalence rate at 3 months. Similarly, as compared with emergency department staff, intensive care unit staff were at intermediate risk, which could be explained by a more usual concern about the risk of pathogen transmission (particularly during highly resistant bacteria outbreaks).

At M0 and M3 and in the analysis restricted to high-risk HCWs and accounting for adherence to PPE measures, being an active smoker reduced the risk of laboratory-confirmed SARS-CoV-2 infection (OR 0.36, 95% CI [0.21; 0.63]). An increasing number of studies have reported this reduced risk of SARS-CoV-2 infection in active smokers in different contexts (cross-sectional studies in the general population; cross-sectional, case–control or control studies in different populations).^17–22^ Our results from a large multicenter prospective study in a young population of HCWs with 26.9% active smokers support the role of use of tobacco substances as protective against SARS-CoV-2 infection, which may act through the nicotine pathway.^23–26^ This result should not encourage smoking to limit the risk of COVID-19; indeed, 78,000 deaths per year are due to smoking in France.^27^ However, the nicotinic hypothesis is of interest, even in the era of an anti-SARS-CoV-2 vaccine, and is being investigating for prevention of COVID-19 (NCT04583410).

The professional category was not a risk factor for laboratory-confirmed SARS-CoV-2 infection, even if medical students exhibited high prevalence as compared with senior physicians (23.5%, 95% CI [20.8-26.4] vs 14.2% [12.2-16.5]) (Table 2). This result may be explained by an insufficient practice or experience with PPE recommendations in medical students or confounded by age. Similarly, reporting high compliance with PPE was not protective (Table 4 and supplementary table 1). However, reported adherence to PPE was very good in the SEROCOV population. Regardless, two studies highlighted greater prevalence of SARS-CoV-2 positive serology in HCWs reporting not systematically wearing PPEs in general or covering the face during clinical encounters than in HCWs fully compliant (15.8% vs 4.3%, *p*= 0.07, and 9% vs 6% *p*=0.012). ^5,6^ The questionnaire on PPE compliance completed only at the end of follow-up in our study may explain in part this discrepancy due to a potential memory bias in addition to social desirability bias.

The strengths of the study are the prospective design, the recruitment in the early phase of the first epidemic wave; the multicenter, multi-department and multi-professional recruitment of the study; the large sample size; the 3-point serology testing; the centralization of serology assays; and the criteria retained for laboratory-confirmed SARS-CoV-2 infection (rather than seroprevalence alone, which is exposed to negativation). As limitations, first, 11.7% HCWs were lost to follow-up at M3, but missing serology results were handled by multiple imputations. Second, compliance with PPE and standard recommendations were queried only at the end of the study and not weekly, as for the clinical signs, with therefore a potential risk of memory bias and social desirability bias. Finally, the serology assay we used (targeting the viral nucleoprotein) is less sensitive than anti-spike assays.^8,12^ Therefore, the reported incidence rate of documented SARS-CoV-2 infection in the SEROCOV study may be slightly underestimated. However, we accounted for this issue in the comparison with seroprevalence data available in the general population.

In summary, in this study of frontline HCWs in Paris, France, we report a 14.6% SARS-CoV-2 cumulative incidence rate at the end of the first COVID-19 wave, with a seroprevalence in May 2020 significantly higher than in the general population. The study of risk factors for laboratory-confirmed SARS-CoV-2 infection argues for a significant part of professional (together with household) contaminations and highlights smoker status as an independent protective factor. At the era of anti-SARS-CoV2 vaccination, our results represent an argument to include HCWs in the first-line target population.

## Data Availability

on request

## Acknowledgements

We sincerely thank all the HCWs for their participation. We specifically thank for their exceptional involvement in data management and collection of blood samples: Ilaria Cherubini, Enfel Houas, Guillaume Payan, Sylvie Le Gac, Odile Fleurot, Elisabeth Bouvet, Christine Jestin, Justine Dorchies, Nathanelle Yeni, Olivia Da Conceicao, Lucie Touchar, Nicolas Mediamolle, Margot Dropy, Aida Zarhrate, Fatiha Bouchama, Marina Vaz, Mariem Ben Cheikh Souguir and Pr Vincent Calvez for virology laboratory staff management. We thank Laura Smales for English editing.

The sponsor of the study was Assistance Publique-Hôpitaux de Paris (AP-HP), with study management by URC Pitié-Salpêtrière. This study was funded by the French Ministry of Health (Programme Hospitalier de Recherche Clinique) and the French Agency for Research (Fond d’amorçage de l’Agence National pour la Recherche).

The sponsor and funders did not participate in the study design, analysis, interpretation of the results or drafting of the manuscript. FT, PH and DH had full access to all data in the study.

## Authors contribution

study design and writing the protocol (PH, FT, LG), participants recruitment and data monitoring (KL, AB, MDu, JMC, JG, AC, NC, RG, MDj, RB, SM, VP, LG, CM, DH), virology tests management (DB,ET, AGM), statistical analysis (DH), analysis of data (all authors) manuscript writing (PH, FT). All authors reviewed the final version of the manuscript.

The study is registered on ClinicalTrials.gov: NCT04304690

## Data sharing statement

Data collected for the study, including individual participant data and a data dictionary defining each field in the set, will be made available to others under request. The study protocol and statistical analysis plan will be available on request on an URL of the URC Pitié Salpêtrière Charles Foix.

